# Assessing the Capacity, Barriers, and Facilitators of the Health Care System to Handle Neglected Tropical Diseases in a Refugee Settlement in Uganda: A Mixed Methods Study

**DOI:** 10.1101/2025.10.16.25338138

**Authors:** Elvis Tamale, Paddy Derrick Malinga, Mable Ssebaduka Nabweteme, Celine Ahurira, Conrad Makai, Kenneth Ssembuze, Patricia Ajeru, Patience Atuhaire

**Author notes:** **Corresponding author**: Tamale Elvis, St. Mary’s Hospital, Lacor, Gulu City, Uganda.

## Abstract

**Background:** Neglected tropical diseases (NTDs) disproportionately affect over one billion people globally, particularly the poorest communities, with Uganda facing a significant burden where over 40 million people are at risk. Despite global efforts by the World Health Organization (WHO) to integrate NTD management into primary health care (PHC) systems, the capacity of health facilities in refugee settings to diagnose and manage these diseases remains largely unknown. This study aimed to assess the capacity of PHC facilities in Nakivale Refugee Settlement, Uganda, with a focus on health workers’ knowledge of NTDs (specifically, soil-transmitted helminths and schistosomiasis) and the perceived facilitators of and barriers to their diagnosis and management.

**Methods:** A mixed-methods cross-sectional study employing quantitative and qualitative methods was conducted. We administered a pre-tested questionnaire to health workers in Health Centre III (HCIII) and Health Centre IV (HCIV) facilities within the Nakivale Refugee Settlement. In-depth interviews were conducted with 22 health workers. A thematic analysis approach was employed to code and categorize the qualitative data systematically. Descriptive statistics, including frequencies, percentages, means, and standard deviations, were used, while qualitative themes were organized according to the research objectives, focusing on health workers’ knowledge of NTD symptoms and transmission, the availability of diagnostic tools and medications, and the systemic and community-level facilitators and barriers to effective NTD diagnosis and management.

**Results:** The study population comprised 150 health workers, predominantly females (60%) with a mean age of 31.5 years, who mostly held diplomas or certificates. Regarding capacity criteria, the findings were striking: 88% of health workers reported no previous training on NTDs, and 84% had not received supportive supervision for NTDs. The process of reporting was also challenging; while 39% found it easy, 38% found it difficult, and 71% reported that the NTD reporting form was difficult to fill out. In terms of knowledge, participants were more aware of the signs of urinary schistosomiasis, with 65% knowing about *S. haematobium*, compared to 58% for *S. mansoni*. The qualitative findings identified key barriers, including a lack of knowledge among health workers and communities, and the systemic under-prioritization of NTDs due to funding and a lack of supplies. Facilitators for improved management included health worker and community education programmes, integrated outreaches and Mass Drug Administration (MDA), and strengthening diagnostic capacity.

**Conclusion:** The findings reveal a critical gap in NTD-specific training and supportive supervision among health workers in the Nakivale refugee settlement. While basic knowledge of common NTD types, symptoms, and medications exists, the lack of formal training and the perceived complexity of reporting systems pose significant barriers to effective NTD management. Targeted interventions focusing on comprehensive training, structured supervision, and simplification of reporting mechanisms are crucial to enhancing the capacity of PHC facilities and facilitating the integration of NTD programs in refugee settings, thereby accelerating the realization of Sustainable Development Goal 3.

## Introduction

Neglected tropical diseases (NTDs) are a group of 13 infections caused by parasitic worms, protozoa, or bacteria currently affecting over 1 billion people, with another 1 billion at risk of infection [1–8, 25–31, 33]. They strike the world’s poorest people with adverse effects on the health, well-being, and socioeconomic facets of one’s life [2, 9–11, 34]. Africa bears a substantial portion of this global burden, accounting for over 600 million cases, particularly affecting poor rural communities [1–2, 12, 36]. Uganda is endemic for all five NTDs, with more than 40 million people at risk for one or more NTDs targeted by the United States Agency for International Development (USAID) Act to End NTDs [13, 37].

Despite their significant burden, NTDs are largely preventable and amenable to elimination. In 2020, the World Health Organization (WHO) outlined a roadmap for eradicating and eliminating 20 NTDs by 2030 [14, 38]. The WHO, alongside policymakers and academics, advocates for integrating NTD management into existing public health programs within endemic countries [39, 40]. Such integration is posited to contribute significantly to achieving several sustainable development goals (SDGs) and vice versa [14–16, 32, 35]. Despite the burden of NTDs in Sub-Saharan Africa, NTDs have not been well integrated into the PHC of most countries. Previous studies in Nigeria, Tanzania, and Burundi have consistently reported low capacity and readiness among health workers to diagnose and manage NTDs [1, 17–18, 41, 42].

Refugee settlements present unique health vulnerabilities that can exacerbate the prevalence of NTDs. Overcrowded living conditions and inadequate sanitation within these camps create conducive environments for the propagation of vectors and parasites associated with diseases such as leishmaniasis and schistosomiasis [20–21]. Furthermore, refugees often encounter significant barriers to accessing healthcare, including logistical, legal, and language challenges, which impede timely medical intervention and increase the burden of NTDs in these vulnerable populations [22].

Nakivale Refugee Settlement, the eighth-largest settlement globally, hosts approximately 145,613 refugees from ten different nationalities [19, 43]. Within this settlement, NTDs, particularly schistosomiasis and soil-transmitted helminths (STHs), are prevalent, with reported rates of 26.6% and 26.5%, respectively [23]. The increasing number of refugees, coupled with inadequate resources, raises critical questions about the capacity of the existing health system to effectively diagnose, manage, and treat these NTDs. Moreover, comprehensive data on NTDs and their prevalence are scarce in regions such as southwestern Uganda. Uganda’s national strategy for NTDs involves integrated control programs [24]. The Ministry of Health has identified several hindrances to the elimination of NTDs by 2030, including a lack of knowledge, skills, and equipment for diagnosis and management in health facilities, as well as deficiencies in reporting systems [23].

Exploring the readiness and capability of PHC facilities to diagnose and manage NTDs is paramount, as it provides a crucial baseline for tracking progress and informing targeted interventions. This study explored the capacity, barriers, and facilitators of PHC centers in Nakivale Refugee Settlement to diagnose and manage soil-transmitted helminths and schistosomiasis.

## Methods

### Study Design and Setting

This study employed a mixed-methods cross-sectional study employing quantitative and qualitative methods. A pre-tested questionnaire was administered to 150 health workers in Health Centre III (HCIII) and Health Centre IV (HCIV) facilities within the Nakivale Refugee Settlement. In-depth interviews were conducted with 22 health workers to assess the capacity of the healthcare system in managing neglected tropical diseases (NTDs) within Nakivale Refugee Settlement, located in southwestern Uganda. Nakivale is one of the largest refugee settlements in the country, hosting a diverse population from over ten countries. The study focused on eight primary healthcare facilities, specifically Health Centre III and Health Centre IV facilities, which provide frontline care to the settlement’s population.

### Study Population and Sampling

The study population comprised health workers actively engaged in service delivery at the selected facilities. For the quantitative study, the initial sample size of 128 participants was determined using the modified Kish equation (44-45) for a general population of 190. A finite population correction was applied to the sample size, and a 10% non-response rate was included, yielding a final sample of 150 participants who were recruited consecutively across the eight facilities. For the qualitative aspect, key informant interviews were conducted with 22 HCWs who had participated in the management of NTDs. The study specifically focused on HCW in the Nakivale Refugee Settlement. Participants were healthcare providers directly involved in the prevention, diagnosis, treatment, or management of NTDs purposively selected with guidance from the District Health Officers (DHOs) and facility in-charges within the settlement. Selection was based on their roles and expertise in NTD management and included HCWs from Health Centers III and IV. The

### Data Collection

Data were collected using a structured self-administered questionnaire adapted from OpenWHO resources on NTDs. The questionnaire collected information on participants’ sociodemographic characteristics, including age, sex, marital status, education level, cadre of health personnel, and the type of health facility in which they worked. Participants were also asked about their knowledge of NTDs, including recognition of schistosomiasis subtypes (S. haematobium and S. mansoni), soil-transmitted helminth subtypes (hookworm, roundworm, whipworm, tapeworm, and flukes), the clinical presentation of these infections, and commonly used medications for their management. Additional questions addressed prior training on NTDs, whether participants had received supportive supervision for NTD management, and their perceptions of NTD reporting systems, including familiarity with reporting forms and views on the role of monetary incentives in promoting reporting.

Interviews were conducted by members of the research team trained in qualitative data collection methods. Each interview was conducted by one member, with the other taking memos. Interviews were conducted from 10 April to 15 May 2025 and lasted 40–60 minutes, with an average duration of 45 minutes. The interviews were conducted in person. All interviews were conducted in the English language, given that HCWs are taught in English. Interviews were audio-recorded and transcribed by two independent members of the research team. The interview guide was generated through reading literature relevant to NTDs in sub-Saharan Africa. The guide included questions focusing on the burden of common NTDs like STHs and schistosomiasis, their causes, transmission modes, management practices, and community impacts to understand the perspectives of the HCW. Additional inquiries focused on challenges faced in NTD management, such as health system issues, diagnostic test availability, drug resistance, and case reporting. The guide also sought suggestions for overcoming these challenges and any additional insights from the interviewees. These were used as probing points to guide the interviews.

### Operational Definition

For this study, neglected tropical diseases (NTDs) were defined as soil-transmitted helminths (STHs) and schistosomiasis.

### Study Variables

Independent variables included sociodemographic characteristics such as age, gender (male, female), education level (certificate, diploma, bachelor’s degree, master’s), cadre or position held at the health facility according to Ministry of Health guidelines, and type of health facility (Health Centre III or IV). Other independent variables included prior training on NTDs, previous training on NTD surveillance and reporting, history of supportive supervision for NTDs, perceptions on whether reporting requires monetary incentives, and whether the NTD reporting form was considered easy to complete. The dependent variable was health workers’ knowledge of the symptoms and management of STHs and schistosomiasis. Knowledge was assessed using ‘yes’ or ‘no’ items, with correct answers scored as 1 and incorrect answers scored as 0. Total scores ranged from 0 to 23 and were categorized as high knowledge (>16, ≥75%), moderate knowledge (12–16, 50–74.9%), or low knowledge (<12, <50%).

### Data Analysis

**Quantitative data** from questionnaires were cleaned in Excel, exported to STATA 17.0, and analyzed using descriptive and inferential statistics. Continuous variables (e.g., age) were summarized with means, medians, standard deviations, and ranges, while categorical variables (e.g., cadre, training, supervision, reporting practices) were described using frequencies and percentages. Multiple-response items were disaggregated and coded. Associations were tested using chi-square or Fisher’s exact tests, with variables significant at p<0.20 included in multivariable logistic regression. Significance was set at p<0.05. Results were presented in tables, charts, and graphs.

**Qualitative data** from interviews were transcribed, anonymized, proofread, and validated through member checking. Thematic analysis followed six phases: familiarization, coding, theme identification, review, definition, and reporting. A codebook was developed in ATLAS.ti, and relational content analysis was applied across transcripts. Weekly team discussions ensured consistency in coding and interpretation. Trustworthiness was enhanced through member checking and triangulation.

### Ethics Statement

This study was conducted in accordance with the ethical principles outlined in the Declaration of Helsinki. Ethics approval was obtained from the Lacor Hospital Institutional Research Ethics Committee (Reference Number: LACOR-2024-362). Written informed consent was obtained from all participants prior to data collection. Participants were informed about the study objectives, procedures, potential risks and benefits, their right to decline or withdraw at any time without any consequence, and confidentiality measures were assured.

## Results

### Demographic characteristics of health workers

The study involved 150 health workers. Table 1 shows the baseline demographic characteristics of the health workers involved in the study. The mean age of the respondents was 31.5 years (SD = 6.41), with ages ranging from 21 to 61 years. A majority of the respondents were female (60%, n=90), whereas 40% (n=60) were male. Most health workers were employed at health center III (HCIII) facilities (74%, n=111), with the remaining 26% (n=39) employed at health center IV (HCIV) facilities.

**Table 1:** Baseline Demographic Characteristics of Health Workers (N=150)

| Characteristic | Category | N | Percentage(%) |
| --- | --- | --- | --- |
| Age(Years) | Mean(SD) | 31.5 (6.41) | N/A |
|  | Range | 21-61 | N/A |
| Gender | Male | 90 | 60 |
|  | Female | 60 | 40 |
| Health Facility Type | Health Center III | 111 | 74 |
|  | Health Center IV | 39 | 26 |
| Education Level | Certificate | 66 | 44 |
|  | Diploma | 69 | 46 |
|  | Degree | 15 | 10 |
| Position Held | Enrolled Nurse | 43 | 29 |
|  | Enrolled midwife | 38 | 25 |
|  | Clinical Officer | 26 | 17 |
|  | Medical Officer | 7 | 5 |
|  | Medical Superintendent | 2 | 1 |
|  | Others | 34 | 23 |

In terms of education, 46% (n=69) held a diploma, 44% (n=66) had a certificate, and 10% (n=15) possessed a degree. The most prevalent positions held were Enrolled Nurse (29%, n=43), Enrolled Midwife (25%, n=38), and ‘Other’ (23%, n=34), followed by Clinical Officer (17%, n=26), Medical Officer (5%, n=7), and Medical Superintendent (1%, n=2).

### NTD Reporting Processes

Table 2 summarizes the perceived challenges in the NTD reporting processes. The perceived ease of reporting on NTDs varied among health workers: 39% (n=58) reported the process ‘Easy,’ 38% (n=57) reported it ‘Difficult,’ 17% (n=25) reported it ‘Very Difficult,’ and only 7% (n=10) reported it ‘Very Easy’. When asked if they found the NTD reporting form easy to fill in, a large majority, 71% (n=107), responded ‘No,’ whereas 29% (n=43) responded ‘Yes’. Regarding the impact of monetary incentives on NTD reporting, opinions were divided: 59% (n=88) believed that they do not promote reporting, whereas 41% (n=62) believed that they do.

**Table 2:** Perceived Challenges in NTD Reporting Processes.

| Characteristic | Category | N | Percentage(%) |
| --- | --- | --- | --- |
| Perceived Ease of Reporting | Very Easy | 10 | 7 |
|  | Easy | 58 | 39 |
|  | Difficult | 57 | 38 |
|  | Very Difficult | 25 | 17 |
| **NTD Reporting Form is Easy to fill** | No | 107 | 71 |
|  | Yes | 43 | 29 |
| **Monetary Incentives Promote Reporting** | Do not promote | 88 | 59 |
|  | Do promote | 62 | 41 |

Health workers reported several deaths attributed to NTDs in recent years, but expressed concern about significant under-reporting in the community. This was often due to delayed diagnoses, refusal of medical treatment, or the preference for religious and traditional healing methods, with symptoms frequently misattributed to non-medical causes like witchcraft (HWs 8, 9, 18). NTD reporting primarily happens through general OPD registers and the HMIS tool (HWs 2, 5, 13). However, the generalized nature of the reporting tools made the identification of the specific STHs and Schistosomiasis difficult. (DHIS tool “should be specific to the type of worms” - HW3).

There was a widespread lack of training on NTD coding among health workers, and overall underreporting due to inactive surveillance. There is less financing and budgeting attached to NTD-related community initiatives (HWs 1, 4, 18). The Ministry of Health’s inconsistency in providing training for health workers about how to fill the reporting forms, the lack of motivation and incentives attached to reporting, was also cited by health workers as a hindrance to reporting (HW 21).

### Training and Supportive Supervision of NTDs

A significant finding was the low prevalence of previous training on NTDs among health workers. Among the 150 respondents, 132 (88%) reported having no previous training on NTDs (soil-transmitted helminths, schistosomiasis), whereas only 18 (12%) had received such training as shown in Figure 1.

**Figure 1:**
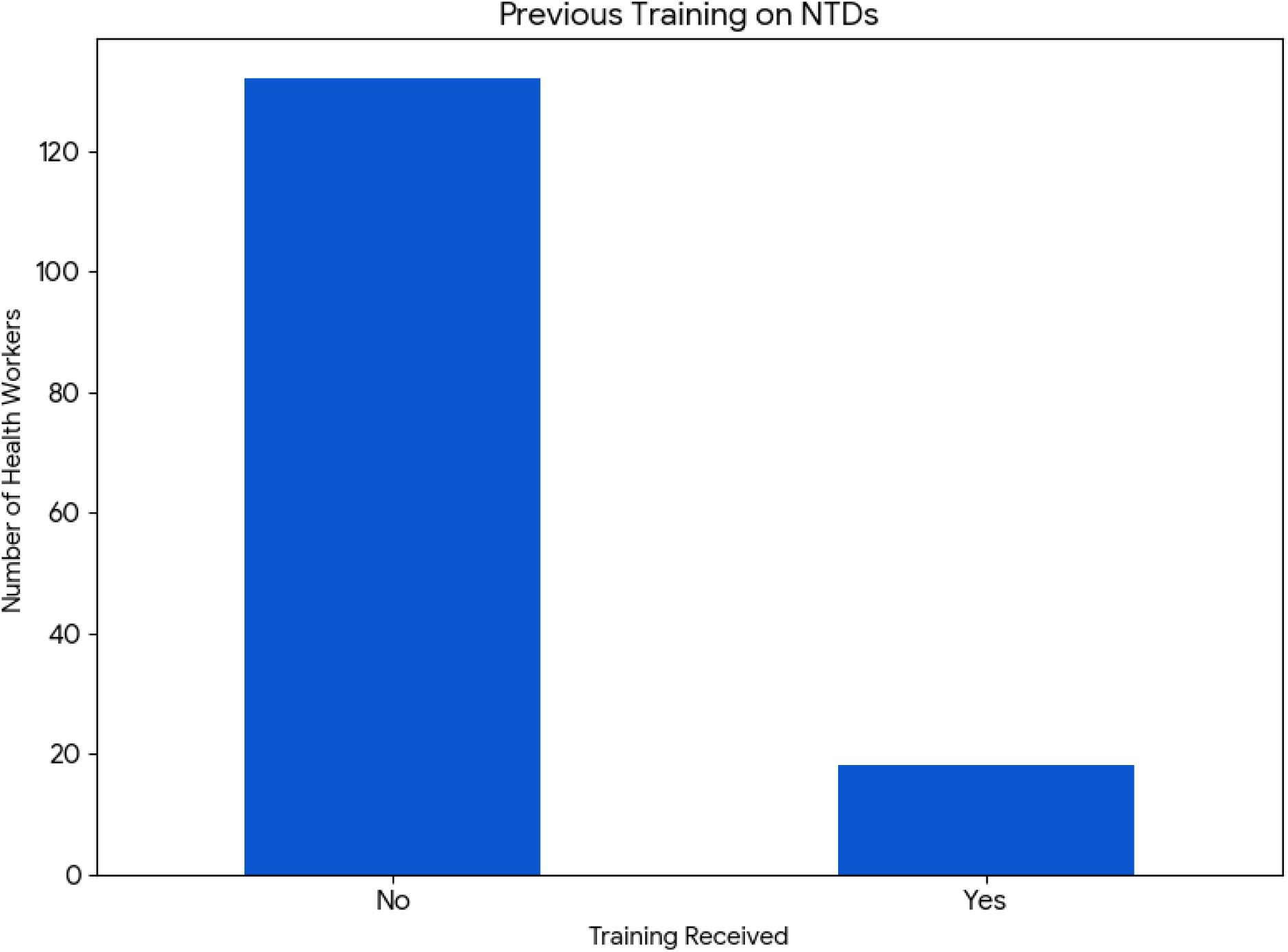
Previous training on NTDs.

Similarly, regarding supportive supervision for NTDs, 126 (84%) health workers reported never having been visited for supportive supervision, whereas only 24 (16%) had received it as shown in Figure 2.

**Figure 2:**
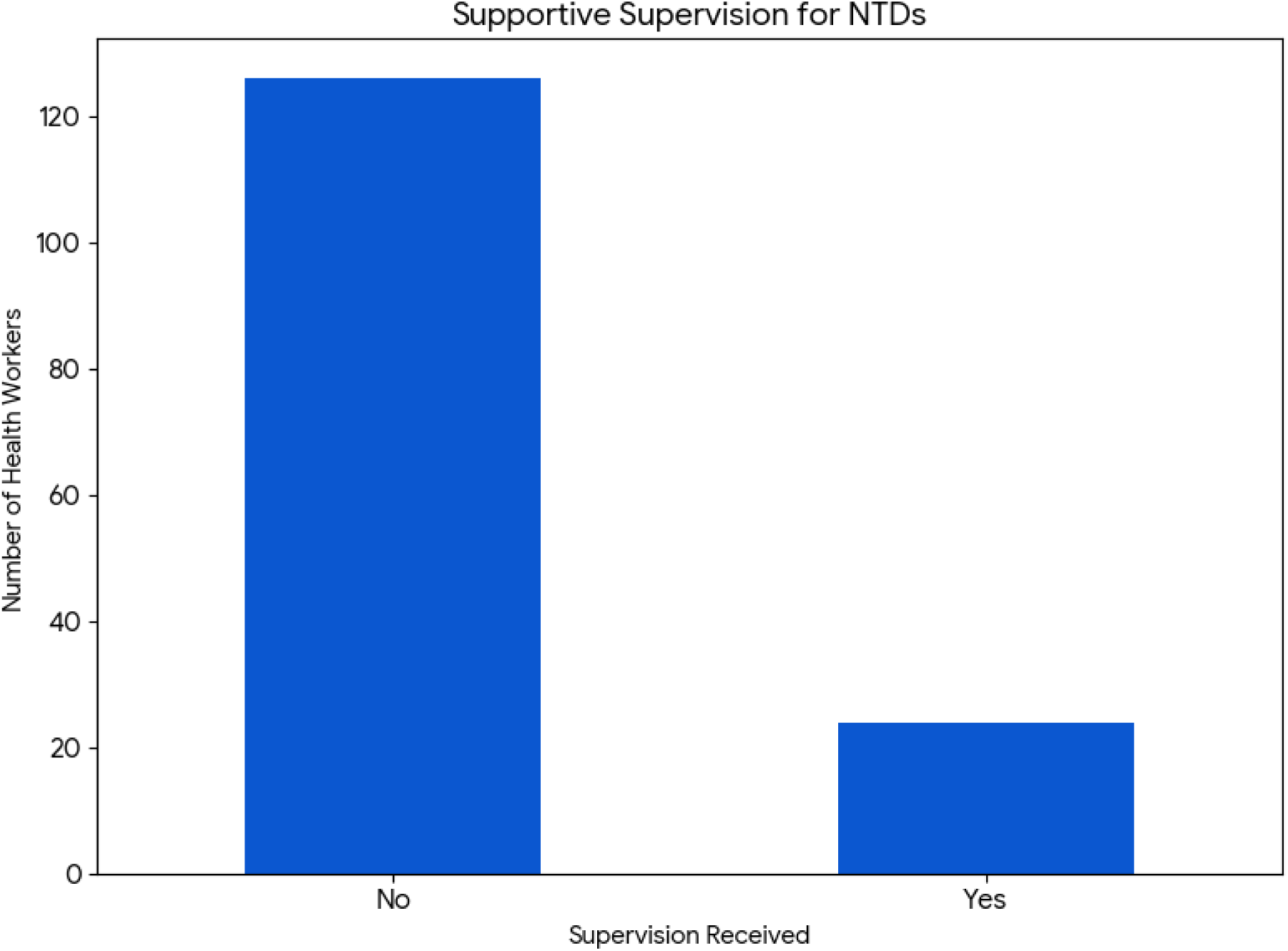
Supportive Supervision for NTDs.

A significant finding was the low prevalence of previous training on NTDs among health workers. Among the 150 respondents, 132 (88%) reported having no previous training on NTDs (soil-transmitted helminths, schistosomiasis), whereas only 18 (12%) had received such training. Similarly, 126 (84%) health workers reported never having been visited for supportive supervision, whereas only 24 (16%) had received it. The Ministry of Health has identified a lack of knowledge, skills, and equipment for diagnosis and management in health facilities as a hindrance to the elimination of NTDs by 2030.

### Knowledge of NTDs

#### Knowledge of NTD Subtypes and Symptoms

Health workers demonstrated a strong understanding of common helminth subtypes, with over 90% awareness for hookworms (95%), roundworms (92%), and tapeworms (91%). Knowledge of schistosomiasis was lower, with 65% of health workers identifying *S. haematobium* and 58% identifying *S. mansoni*. The overall knowledge regarding symptoms of S. *mansoni* (58%) was found to be lower than that for *S. haematobium* (65%) in our study. For *S. haematobium*, anemia and hematuria emerged as the most frequently acknowledged symptoms, at 83% and 81%, respectively. Common symptoms of *S. mansoni* were abdominal pain (83%), anemia (75%), and diarrhea (75%). Our study also revealed a high level of awareness about STH symptoms among health workers, with hookworm (95%), roundworms (92%), and tapeworms (91%) being among the major STHs. The most predominantly identified symptoms were abdominal pain (97%), anemia (93%), and diarrhea (84%).

### Medication Knowledge

The health workers’ knowledge of standard NTD medications was high. An impressive 97% correctly identified albendazole or mebendazole as treatments for STHs. A large majority (74%) knew praziquantel is used for schistosomiasis, and over half (55%) were aware of ivermectin.

From the interviews, the level of knowledge among health workers varied, with at least 15 acknowledging significant gaps and a consensus that training and refresher courses on NTDs are lacking. One health worker, HW14, stated, “We have concentrated on malaria, HIV, and TB, and we have neglected those diseases”. This focus on other priorities, combined with the non-specific symptoms of many NTDs, means that the diseases often receive less clinical attention. HW21 noted a crucial point: “Once you sit in that clinic, usually what you don’t know, you’re less likely to think about it”.

Concerning NTD symptoms and transmission methods, there were key knowledge gaps. Health workers did, however, identify common symptoms such as “abdominal pain and some bit of diarrhea, blood in stool” (HW22) for helminths, and “lower back pain… painful urination… hematuria” (HW14) for schistosomiasis. Lake Nakivale was perceived as a primary transmission source by the majority of participants, especially at lakeshores and landing sites, where activities like swimming and fishing were frequently associated with higher risks of contracting NTDs. Fecal–oral transmission, closely connected to the lack of proper sanitation and hygiene practices, was identified as another significant pathway for disease spread. Cultural beliefs against using latrines and the absence of handwashing facilities exacerbated the situation. Poverty was also noted to be a major contributing factor to NTD exposure, with many children going to school without shoes, making them vulnerable to helminth infections.

Health workers reported varied insights into the NTD burden, with many linking a perceived low burden to diagnostic limitations and low community awareness. One health worker, HW1, stated, “It may be a burden that is not discovered because of the knowledge gaps.” Another, HW21, added, “You can’t tell the burden of something that you don’t know”. Intestinal helminths (hookworms, roundworms, tapeworms) and schistosomiasis were the most frequently reported NTDs, with one frontline respondent putting cases at “roughly like 10 intestinal worm cases a month”.

The most affected groups identified were children and teenagers, particularly those in school, as well as fishermen who are susceptible due to their proximity to Lake Nakivale, which is a community water source where a lot of waste is dumped. HW22 specified, “Most of the people that we have diagnosed are teenagers, and the majority are between the ages of 10 to 18”. HW9 noted, “Of the two cases so far I know, all of them are fishermen,” directly linking their economic activity to exposure. Other NTDs, such as elephantiasis, trachoma, tungiasis (jiggers), and leprosy, were reported to be less frequent.

Health workers also noted the impact of NTDs on productivity, with one stating that “a sick person is not productive” (HW21). The burden was also connected to seasonal conditions, with stagnant water during wet seasons and shared water sources in dry spells affecting prevalence. Participants emphasized the vital role of interventions initiated by the Ministry and other health agencies in controlling the spread and severity of these diseases, and believed that community response to sensitization programs and the adoption of better hygiene practices were crucial factors in reducing NTD cases.

### Facilitators and Barriers to Diagnosis and Management Barriers

There was a frequent lack of essential NTD medications and diagnostic tools. Health facilities often face shortages of key supplies like Kato-Katz kits, reagents, and even basic stool containers, as highlighted by one health worker: “Here we are not doing stool, meaning we are missing out on them” (HW14). This forces diagnoses to be based on clinical presentation alone.

From a systemic viewpoint, the study revealed a consistent under-prioritization of NTDs in terms of funding and support, as reflected in their limited budget allocations and the overshadowing of NTD initiatives by other health programs focused on other diseases like HIV and malaria, receiving more funding and attention. This underinvestment leads to a general lack of motivation among health workers to engage with NTD cases, resulting in overlooked diagnoses. One health worker’s comment explicitly summed it up: “Motivation, I can say it is zero per cent” (HW19).

There are also significant knowledge gaps among health workers. Clinicians admitted they often misdiagnose NTDs because they’re not trained to look for them. Clinicians often misdiagnosed NTDs as common conditions like malaria or ulcers, owing to difficulties in suspecting and correctly identifying these diseases. “The challenge is that most of the clinicians are not suspecting, they are just looking at these other signs of malaria and maybe someone is complaining of abdominal pain and whatever and they could say that’s ulcers so when you look at our identification rate is very low, what you don’t know, you’re less likely to think about it” (HW21).

Many community members attribute NTD symptoms to non-medical causes like witchcraft, especially symptoms like an enlarged stomach. This leads them to seek help from traditional healers first. As a result, patients often arrive at health facilities only after traditional remedies have failed, by which point their condition has worsened and is more difficult to treat. “Poor health-seeking behavior, a strong belief in traditional medicines as they usually first try their herbs” (HW21), cultural myths (e.g., believing an illness is “bewitched”, and a lack of community awareness all hinder effective management (HW 3).

Furthermore, there is a widespread lack of community awareness about NTDs. One health worker noted that NTDs are “not something that we always talk about when we are giving health talks” (HW21). This absence of continuous sensitization and information campaigns, like radio shows, hinders efforts to dispel myths and promote timely treatment. These cultural and awareness barriers, combined with financial constraints and poor health-seeking behavior, significantly delay proper diagnosis and care.

### Facilitators and Interventions

Most of the community education programmes focused on infectious diseases such as malaria, HIV, as well as maternal health, with limited focus on NTDs. Health workers suggested empowering communities with knowledge to promote prevention and health-seeking behavior. Utilizing always existent avenues through which communities gain information, such as VHTs, Radio talks, and community outreaches.

HW21 stated, NTDs are not something that we always talk about when we are giving health talks yet it is something we can prioritize in all the avenues we do community education, such as VHTs, radios, and outreaches.“

There should be a focus on integrated Outreaches and Mass Drug Administration (MDA), which includes regular deworming programs and integrated weekly outreaches to educate communities, administer drugs in schools, markets, and areas of convergence, were identified as effective existing interventions (HW 6)

There is a need to organize refresher training and mentorship for health workers to ensure they can improve their “index of suspicion” for NTDs, as well as make a correct diagnosis and management (HW21).

To address the systemic challenges, there should be a focus on strengthening diagnostic capacity, improving WASH infrastructure, and advocating for increased budgets and the inclusion of NTD drugs on essential medicine lists (HW 17).

## Discussion

This study aimed to quantitatively and qualitatively assess the capacity of primary healthcare centers in Nakivale Refugee Settlement, Uganda, for diagnosing and managing soil-transmitted helminths (STHs) and schistosomiasis. It explored health workers’ knowledge of symptoms and signs, availability of diagnostics, as well as barriers to successfully diagnosing and managing STHs and schistosomiasis. Our findings highlight critical gaps related to health worker training, knowledge reinforcement, and reporting systems. The study also offers data-driven insights into the current state of Neglected Tropical Disease (NTD) management in this vulnerable setting.

Neglected Tropical Diseases (NTDs) such as schistosomiasis and intestinal helminths are common in the Nakivale Refugee Settlement, mirroring trends in other refugee and lakeshore communities in Uganda. [22–23]. [21–22, 25]. Children, teenagers, and fishermen are most affected due to frequent contact with Lake Nakivale, poor sanitation, open defecation, and lack of footwear—conditions worsened by poverty and overcrowding. These findings are consistent with studies from Uganda, Ethiopia, and Bangladesh.[20, 36] Health workers demonstrated good knowledge of NTD diagnosis and treatment, especially for schistosomiasis and soil-transmitted helminths, though awareness of *S. mansoni* symptoms was lower than that of *S. haematobium* and lower than reported in Tanzania and Nigeria, suggesting gaps in disease-specific training. [42]. [17–18].

Our study identified major gaps in NTD training and reporting, with only 12% of participants having received prior training—higher than rates in Burundi but far below those in Nigeria. Most health workers (88%) had never received formal instruction, relying instead on informal learning, which contributes to inconsistent diagnosis, treatment, and reporting. Training in NTD surveillance and reporting was also limited (16%), with few participants receiving supportive supervision. Although nearly half found reporting forms easy to use, many described challenges related to unclear procedures, lack of Ministry of Health support, poor incentives, and inactive surveillance systems—highlighting a critical need for structured capacity building and continuous professional development.

Our study identified major gaps in NTD training and reporting, with only 12% of participants having received prior training—higher than rates in Burundi but far below those in Nigeria. [17[1]. Most health workers (88%) had never received formal instruction, relying instead on informal learning, which contributes to inconsistent diagnosis, treatment, and reporting. [1, 17–18, 41, 42 Training in NTD surveillance and reporting was also limited (16%), with few participants receiving supportive supervision.

Although nearly half found reporting forms easy to use, many described challenges related to unclear procedures, lack of Ministry of Health support, poor incentives, and inactive surveillance systems— highlighting a critical need for structured capacity building and continuous professional development.

A shortage of diagnostic and reporting supplies perpetuates a cycle of underdiagnosis and underreporting, leading to underestimated resource needs and limited allocations. [45]. Differences from the Nigerian study likely reflect contextual factors: Nigeria’s urban and semi-urban private health facilities emphasize business-driven efficiency, regular training, and stricter reporting, while our rural, public-sector setting faces weaker systems and lower community awareness. Despite this, our participants had relatively higher training in schistosomiasis and STHs due to hands-on patient care. Strengthening surveillance may require simplifying reporting tools, adopting digital systems, and enhancing non-monetary incentives such as recognition, supervision, and refresher training. Encouragingly, health workers demonstrated strong awareness of STHs—particularly hookworm (95%), ascaris (92%), and tapeworm (91%)—and recognized key symptoms like abdominal pain, anemia, and diarrhea. Children were most affected, often presenting with malnutrition and stunted growth, consistent with findings linking ascaris to malnutrition and hookworm to anemia. The high knowledge levels observed likely reflect the endemic nature of these infections in rural, lakeshore, and refugee settlement contexts. [1]

Our study also revealed an impressive level of awareness about STH symptoms among our respondent health workers, with hookworm (95%), ascaris (92%), and tapeworm (91%) being among the major STHs. The healthcare workers predominantly identified abdominal pain (97%), anaemia (93%), and diarrhea (84%) as symptoms. Children, who were mostly affected by STHs, frequently presented with abdominal swelling, stunted growth, malnutrition, and anemia. This was consistent with other studies that reported ascaris as the main cause of malnutrition among NTDs in children as well as hookworms as the main cause of anemia [44–45]. This discrepancy in knowledge levels compared to other studies could be partly explained by the fact that our study was conducted in rural areas located in refugee settlements and near a lakeshore, which are associated with a higher prevalence of these diseases.

In addition, our study revealed that while most facilities had basic lab infrastructure like microscopes and dyes, critical supply gaps were highlighted, with approximately 90% of respondents highlighting missing essential items like Kato-Katz kits, reagents, or stool containers. This deficiency was mainly attributed to inconsistency in the supply from the government, which forces diagnoses to be based on clinical presentation alone, as HW14 stated, “Here we are not doing stool, meaning we are missing out on them.” The Ministry of Health has also previously cited a general lack of equipment as a hindrance to NTD elimination [23]. The availability of essential medications to treat STHs and schistosomiasis was also a major challenge. The only consistently available drugs were albendazole and mebendazole, with other essential NTD drugs like praziquantel and ivermectin rarely seen, and stock-outs reported. The frequent lack of common medications like praziquantel, which hinders appropriate treatment, was another management difficulty that deters the community from seeking assistance within the facilities. This issue is compounded by a general lack of motivation and training, which contributes to a low “index of suspicion” for NTDs.

Our findings on barriers highlight that the management of NTDs is undermined by a combination of systemic, clinical, and community-level barriers. Health facilities face chronic shortages of essential diagnostic tools and medications, forcing clinicians to rely on clinical judgment alone, which is often inaccurate. The persistent under-prioritization of NTDs in health budgets compared to other conditions such as HIV and malaria not only restricts resources but also dampens health worker motivation.

Additionally, gaps in clinical knowledge and training contribute to frequent misdiagnosis, with NTDs often mistaken for more common conditions. On the community side, cultural beliefs, reliance on traditional healers, and poor health-seeking behaviors delay timely treatment and exacerbate disease progression. The lack of sustained awareness campaigns further entrenches myths and misinformation, leaving both health workers and communities ill-equipped to recognize and address NTDs effectively. Collectively, these factors create a cycle of neglect, where NTDs remain overlooked and under-treated, perpetuating poor health outcomes.

Our findings point to several facilitators that can strengthen NTD prevention and control. Health workers emphasized that existing community education channels—such as Village Health Teams (VHTs), radio programs, and outreach activities—can be leveraged to integrate NTD awareness into ongoing health promotion efforts, thereby improving knowledge and encouraging timely care-seeking[46, 47]. Integrated approaches, including Mass Drug Administration (MDA), regular deworming, and outreach in schools, markets, and other community gathering points, were highlighted as effective platforms to reach large populations with both education and treatment. At the health system level, refresher trainings and mentorship opportunities for clinicians were seen as critical to improve their diagnostic skills and increase their suspicion index for NTDs. In addition, strengthening diagnostic capacity, investing in WASH infrastructure, advocating for dedicated budgets, and ensuring NTD drugs are included on essential medicines lists were identified as structural facilitators that could enhance both case detection and management. Together, these strategies offer a pathway to build on existing systems and resources to improve NTD control.

### Strengths and Limitations

This study provides an assessment of critical capacity aspects within a specific, vulnerable refugee settlement, offering data-driven insights into health worker knowledge, training, and reporting systems for NTDs. It identifies specific numerical gaps that can inform targeted interventions. However, the study has several limitations. First, it relies on self-reported data for knowledge and perceptions, which may introduce recall or social desirability bias. Second, the questionnaire did not directly assess the physical availability of diagnostic tools and specific medications, limiting the depth of analysis for resource capacity. Third, as a cross-sectional study, it offers a snapshot rather than capturing changes over time.

### Recommendations

To enhance the capacity of primary healthcare centers in refugee settings and accelerate progress toward NTD elimination goals, we recommend implementing targeted interventions. These should include the development and delivery of comprehensive and regular NTD-specific training programmes for health workers. Additionally, establishing robust, supportive supervision mechanisms is crucial for continuous professional development and skill reinforcement. Finally, there is a clear need to simplify NTD reporting forms and processes, potentially through digital solutions or more user-friendly designs, to improve data accuracy and completeness.

## Conclusion

The healthcare system in Nakivale Refugee Settlement faces significant challenges in its capacity to handle neglected tropical diseases. While health workers possess foundational knowledge of common NTD types, symptoms, and medications, this knowledge is largely unsupported by formal training and consistent supportive supervision. The current NTD reporting system is perceived as complex and difficult to use, posing a substantial barrier to effective surveillance. Addressing these systemic gaps will be crucial for improving NTD diagnosis, treatment, and management, ultimately enhancing the health and well-being of refugee populations.

## Data Availability

The datasets generated and/or analyzed during the current study are not publicly available owing to the sensitive nature of participant data and to maintain participant anonymity but are available from the corresponding author upon reasonable request.

## List of abbreviations

NTDs: Neglected Tropical Diseases
PHC: Primary healthcare
WHO: World Health Organization
HCIII: Health Centre III
HCIV: Health Centre IV
SD: Standard deviation
STH: Soil-Transmitted Helminths
SDG: Sustainable Development Goal
MOE: Margin of Error
USAID: United States Agency for International Development

## Declarations

### Ethics approval and consent to participate

Following the Declaration of Helsinki, Ethics approval for this study was obtained from the Lacor Hospital Institutional Research Ethics Committee, Reference Number [LACOR-2024-362]. Informed consent was obtained from all participants before data collection.

### Consent for publication

Not applicable

### Authors’ contributions

TEs conceived the study, participated in its design and coordination, and helped draft the manuscript. AP and MPD participated in the study design, performed the statistical analysis, and drafted the manuscript. NMS, AC, MC, SK, AP, and MPD participated in data collection and interpretation. All the authors have read and approved the final manuscript.

## Acknowledgments

The authors would like to thank the health workers in Nakivale Refugee Settlement for their participation in this study. We also extend our gratitude to Medical Teams International for their assistance during data collection. We also extend gratitude to the RSTMH early career grant funding, which facilitated the research process.

